# Blood Eosinophil Reference Values and Determinants in a Representative Adult Population

**DOI:** 10.1101/2024.10.10.24315149

**Authors:** Reshed Abohalaka, Selin Ercan, Lauri Lehtimäki, Saliha Selin Özuygur Ermis, Daniil Lisik, Muwada Bashir Awad Bashir, Radhika Jadhav, Linda Ekerljung, Göran Wennergren, Jan Lötvall, Teet Pullerits, Helena Backman, Madeleine Rådinger, Bright I. Nwaru, Hannu Kankaanranta

## Abstract

**Background:** The use of blood eosinophil count (BEC) as a prognostic biomarker in the management of conditions such as asthma and chronic obstructive pulmonary disease (COPD) may be complicated by factors like atopy, age, sex, smoking, and comorbidities. Therefore, we aimed to produce reference values for BEC, considering age, asthma, COPD, and clinical allergy for the general adult population.

**Methods:** The West Sweden Asthma Study constitutes a population-representative clinical epidemiological cohort of randomly selected adults in western Sweden. From this cohort, *n*=1,145 took part in clinical examinations, including e.g. skin prick testing, specific immunoglobulin E, and BEC.

**Results:** The upper limit (95^th^ percentile) of BEC varied by age. It ranged between 400 to 500 cells/μL in the full sample, and from 300 to 400 cells/μL in subjects without asthma, COPD, and clinical allergy (*n*=710). Sex, smoking, atopy, clinical allergy, obesity, asthma, COPD, diabetes, and hypertension were statistically significantly associated with higher BEC levels. However, only asthma and clinical allergy in the full sample, and obesity and diabetes in those without asthma, COPD, or clinical allergy, remained statistically significant in multiple regression analyses.

**Conclusion:** In a population-representative sample, the upper limit of BEC in healthy adults ranged between 300 and 400 cells/μL, varying by age. Age, smoking, obesity, asthma, COPD, and clinical allergy influence BEC levels and should be considered in clinical interpretation.

**Clinical Implications:** The upper limit of normal blood eosinophil count (BEC) in participants free from asthma, COPD, and clinical allergy ranged between 300 and 400 cells/μL depending on age. These results facilitate the interpretation of BEC in clinical practice.

**Capsule summary:** The upper limit of normal blood eosinophil count (BEC) in healthy individuals ranged between 300 and 400 cells/μL depending on age. Determinants of BEC values include age, smoking, obesity, asthma, and clinical allergy.

**Support statement:** The study was supported by the VBG Group Herman Krefting Foundation for Asthma and Allergy Research (Trollhättan, Sweden), Swedish Research Council (Stockholm, Sweden), the Swedish Heart-Lung Foundation (Stockholm, Sweden), the Swedish Asthma and Allergy Foundation (Stockholm, Sweden), Tampere Tuberculosis Foundation (Tampere, Finland), and ALF agreement (grant from the Swedish state under the agreement between the Swedish Government and the county councils, Sweden).

## Introduction

Blood eosinophil count (BEC) plays a pivotal role in determining the severity, phenotypes, and therapy of a spectrum of chronic inflammatory lung disorders, most notably asthma and chronic obstructive pulmonary disease (COPD) [1,2]. Indeed, BEC is a valuable prognostic indicator for assessing responsiveness to anti-interleukin 5 (anti-IL5) and anti-IL-4/13 therapies in severe asthma [3], as well as for predicting the efficacy of inhaled corticosteroids (ICS) in COPD [4,5]. In addition, the Global Initiative for Asthma (GINA) recommends the use of BEC to identify asthma patients with type 2 (T2) inflammation [2]. Meanwhile, the Global Strategy for the Diagnosis, Management and Prevention of Chronic Obstructive Lung Disease (GOLD) recommends the use of 300 cells/μL as a threshold of circulating eosinophils to guide therapy with ICS in patients with COPD who continue to exacerbate despite appropriate bronchodilator therapy [1].

However, the application of BEC as a biomarker in clinical practice is not straightforward. First, the predictive cut-off points for BEC interpretation derived from randomized controlled trials are confined to studies conducted within asthma and COPD populations [6–16]. Second, the recommended BEC thresholds in clinical guidelines are subject to debate due to their perceived overlap with normal eosinophil ranges [17,18]. This controversy arises from the limited evidence of normal and abnormal blood eosinophil levels across diverse populations with varying backgrounds and health conditions. This evidence gap is highlighted by the fact that investigations into healthy populations suggest that BEC are influenced by several demographic and clinical factors, including age, sex, obesity, atopy, and smoking [19–21].

Furthermore, studies having representative random samples from the general population are scarce. A recent systematic review by *Benson et al.* [21] that aimed to synthesize the absolute BEC rather than reference values, identified 14 studies conducted in the general population.

Only one of these studies used a random sample directly from the population itself [22]. Conversely, two additional studies, while encompassing a considerable sample size, did not recruit participants from the general population [20] nor did report reference values for the healthy cohort [23]. Thus, the typical range of BEC in the general population is not well-established. Therefore, our objective was to determine the BEC reference values and its determinants in a population-representative sample.

## Methods

### Study area and population

The West Sweden Asthma Study (WSAS) has been described in detail previously [24]. Briefly, WSAS examines individuals aged 16 to 75 years at recruitment, randomly selected from the general population of western Sweden. Commencing in 2008, a total of 30,000 subjects within the specified age range were randomly selected through the Swedish Population Register and invited to take part in a postal survey. Of those invited, 18,087 individuals participated in the survey study. Subsequently, a random subset of 2,000 individuals were invited to comprehensive clinical examinations, of which 1,145 participated (**Figure 1**). All participants signed informed consent, and the study was approved by the regional ethics board in Gothenburg, Sweden.

**Figure 1.**
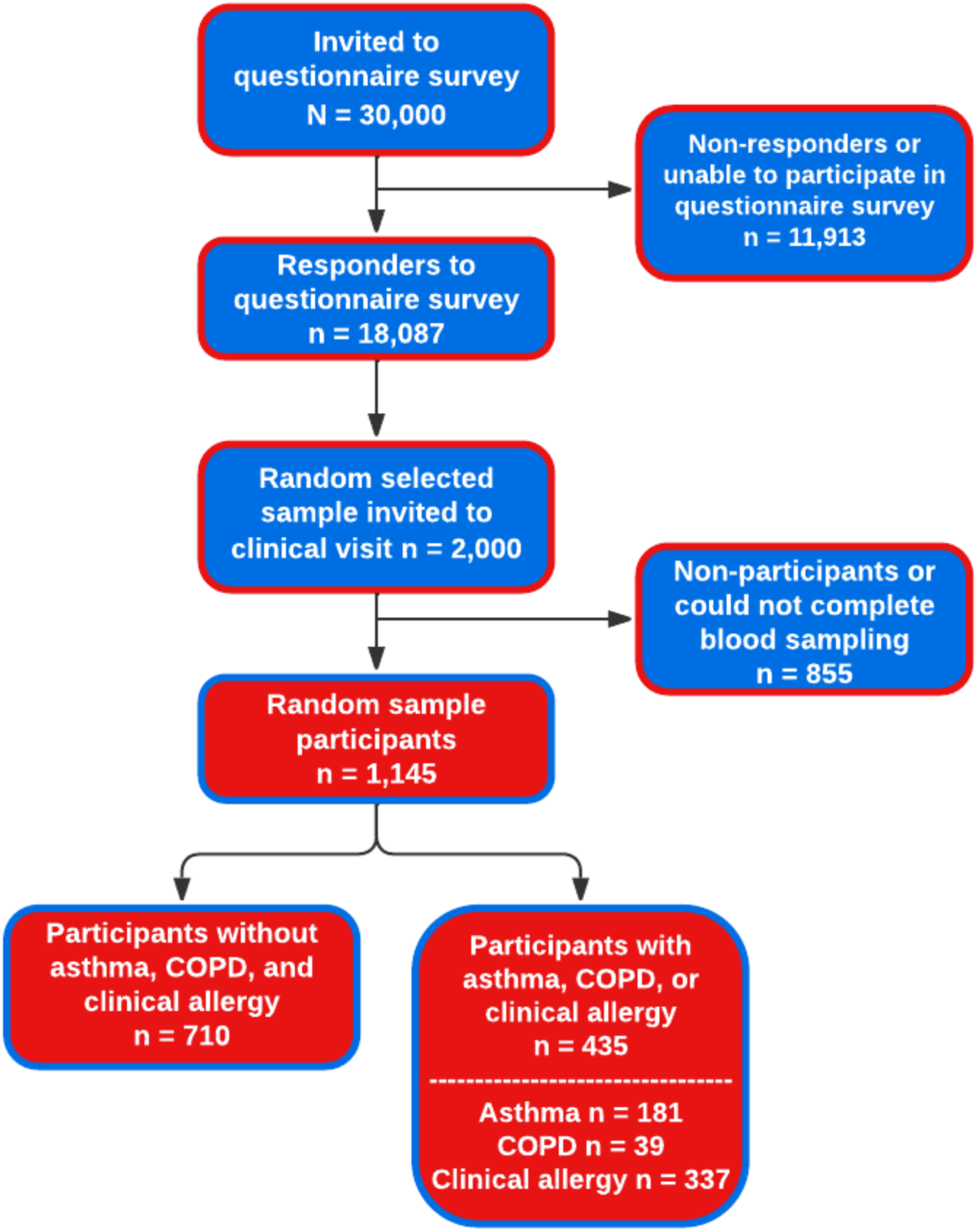
Flow chart of participation in the study. Clinical allergy was defined by the presence of allergic sensitization coupled with consistent self-reported allergic symptoms attributable to the same allergen. COPD: Chronic Obstructive Pulmonary Disease.

### Clinical examinations

The examinations included, but not limited to, blood cell quantification, skin prick testing (SPT), specific immunoglobulin E (sIgE) level assessment, and measurement of height and weight. Additionally, the clinical examination encompassed in-depth, structured interviews and administration of questionnaires pertaining to respiratory diseases and symptoms, morbidities, healthcare utilization, and potential risk factors.

### Assessment of sensitization and blood eosinophils

Sensitization was evaluated through the determination of sIgE levels and SPT for 11 aeroallergens (Details are provided in the **supplementary file**). BECs were determined using standard procedures at Sahlgrenska University Hospital (Gothenburg, Sweden) with ADVIA® 2120i Hematology System (Siemens Healthineers, Erlangen, Germany), and are reported as the number of cells per microliter (μL).

### Definitions of diseases

Clinical allergy was defined as the presence of allergic sensitization, indicated by either a positive SPT or elevated sIgE level to at least one allergen (atopy), coupled with self-reported allergic symptoms attributable to the same allergen. The presence of metabolic disorder was confirmed by obesity (body mass index [BMI] ≥30 kg/m^2^) or any of the following self-reported conditions: hypertension, hyperlipidaemia, or diabetes [25]. Detailed definition of Asthma and COPD are provided in the **supplementary file.**

### Statistical analyses

In the establishment of reference values, the presumption is that the spectrum of values derived from a cohort of healthy individuals is a proxy for normal values. Consequently, individuals manifesting values beyond the norm, typically delineated by the central 90% of the range of values in the healthy population (upper limit of normal or 95^th^ percentile and lower limit of normal or 5^th^ percentile), are frequently characterized as exhibiting anomalous outcomes [26, 27]. As a result, our reporting considers the 95^th^ percentile of BEC as the upper limit of normal. Due to the non-normal distribution of BEC values exhibiting a right-skewed shape, percentiles were derived from the logarithmically transformed dataset, as previously described [20]. Statistical comparisons were carried out using independent Student’s t-tests. Statistical significance was deemed present at a threshold of *p*<0.05.

A multiple binary logistic regression model was applied to compute odds ratios (OR) accompanied by 95% confidence intervals (CI) for the purpose of elucidating associations between various factors and BEC values higher than the upper limit (95^th^ percentile), adjusting for potential confounding variables. All statistical analyses were executed utilizing SPSS 29.0 (IBM Corp, New York, USA).

## Results

### Characteristics of the random sample

The random sample comprised of 1,145 individuals, with 46.8% being males. The mean age of the total study sample was 50.4±15.4 years (mean±standard deviation (SD)). Nearly half of the participants (50.8%,*n*=582) reported no history of smoking, while 11.4% were current smokers. Of the participants, 37.8% *(n*=426) were sensitized (atopic), while 29.8% *(n*=337) met the criteria for clinical allergy (**Table 1**). Individuals with clinical allergy were younger (45.6±14.7 years), had more males, and more non-smokers than those without clinical allergy (**Table 1**). For other characteristics of those with asthma with or without clinical allergy, see **Table S1**.

**Table 1.** Characteristics of study subjects with or without asthma or clinical allergy (N=1,145).

|  | Without<br>Asthma | With Asthma | Without<br>Clinical allergy | With Clinical<br>allergy |
| --- | --- | --- | --- | --- |
| Subjects n (%) | 964 (84.2) | 181 (15.8) | 792 (70.2) | 337 (29.8) |
| Asthma n (%) | - | 181 (100) | 65 (8.2) | 115 (34.1) |
| Clinical allergy* n (%) | 222 (23.4) | 115 (63.9) | - | 337 (100) |
| **Demographics** |  |  |  |  |
| Age (y) mean (SD) | 50.7 (15.5) | 48.7 (15.1) | 52.1 (15.3) | 45.6 (14.7) |
| Male sex n (%) | 456 (47.3) | 80 (44.2) | 349 (44.1) | 179 (53.1) |
| BMI, kg/m<sup>2</sup> mean (SD) | 26.0 (4.1) | 26.9 (4.5) | 26.1 (4.1) | 26.2 (4.3) |
| Education years over 12 years n (%) | 437 (45.4) | 90 (50.0) | 345 (43.6) | 175 (52.9) |
| Never smokers n (%) | 496 (51.5) | 86 (47.5) | 380 (48.0) | 196 (58.2) |
| Current smokers n (%) | 103 (10.7) | 27 (14.9) | 99 (12.5) | 29 (8.6) |
| Pack-year mean (SD) | 14.0 (12.8) | 17.5 (15.9) | 14.8 (13.1) | 13.3 (14.0) |
| **Allergic symptoms** |  |  |  |  |
| Age at onset of symptoms mean (SD) | 24.5 (16.8) | 17.8 (15.0) | 28.6 (18.1) | 18.6 (14.3) |
| Eyes and nasal symptoms n (%) | 249 (49.2) | 106 (68.8) | 119 (33.1) | 234 (79.9) |
| Ever allergic nasal symptoms n (%) | 301 (31.3) | 129 (71.3) | 152 (19.3) | 273 (81.0) |
| Current allergic nasal symptoms n (%) | 454 (88.0) | 147 (94.8) | 309 (84.5) | 285 (96.0) |
| **Atopy and allergy** |  |  |  |  |
| ≥1 positive reaction in SPT# n (%) | 223 (34.0) | 102 (72.9) | 60 (11.7) | 265 (94.0) |
| ≥1 positive sIgE n (%) | 251 (27.4) | 111 (63.4) | 65 (8.5) | 297 (91.7) |
| Atopic (SPT or sIgE) n (%) | 304 (32.1) | 122 (67.8) | 92 (11.6) | 337 (100) |
| Clinical allergy to mite n (%) | 42 (4.4) | 42 (23.3) | - | 84 (24.9) |
| Clinical allergy to furry animals n (%) | 74 (7.8) | 71 (39.4) | - | 145 (43.0) |
| Clinical allergy to pollen n (%) | 168 (17.7) | 97 (53.9) | - | 265 (78.6) |
| Clinical allergy to nuts n (%) | 40 (4.4) | 49 (28.0) | - | 89 (27.5) |
| Clinical allergy to seafood or dairy n (%) | 38 (4.1) | 19 (10.9) | - | 57 (17.6) |
| **Morbidities** |  |  |  |  |
| COPD | 22 (2.3) | 17 (9.4) | 26 (3.3) | 10 (3.0) |
| Obesity (BMI ≥ 30 kg/m<sup>2</sup>) | 132 (13.7) | 40 (22.1) | 113 (14.3) | 55 (16.3) |
| Diabetes mellitus | 37 (3.8) | 9 (5.0) | 27 (3.4) | 18 (5.4) |
| Hypertension | 225 (23.5) | 48 (26.7) | 197 (25.0) | 67 (20.1) |
| Other cardiovascular diseases | 113 (11.7) | 23 (12.7) | 98 (12.4) | 32 (9.5) |
| Hyperlipidaemia | 151 (15.7) | 32 (17.7) | 129 (16.3) | 47 (13.9) |
| Metabolic disease | 377 (39.1) | 83 (45.9) | 321 (40.5) | 127 (37.7) |
| Gastroesophageal reflux disease | 409 (42.5) | 102 (56.4) | 342 (43.3) | 163 (48.4) |

Data was presented as n (%) or mean (SD). BMI= body mass index, SPT = skin prick test, sIgE = specific immunoglobulin E, COPD = chronic obstructive pulmonary disease. \* Some participants (N = 16) did not undergo SPT test nor sIgE, so they have been excluded from the total count of clinical allergy. #Some participants (N = 349) did not undergo SPT test, so they have been excluded from the total percentage.

### Blood-eosinophil count in the random sample

In the full random sample, the upper limit of BEC was 400 cells/μL across all ages, except for those aged 50-60 years, where it reached nearly 500 cells/μL (**Figure 2A**). To assess the influence of asthma and COPD on BEC, we excluded patients with these conditions (**Figure 2B**). The 95^th^ percentile BEC remained stable at 400 cells/μL for those without asthma and non-COPD individuals across different ages, except for those aged 30-40 years, where it was around 300 cells/μL. Next, we examined the impact of clinical allergy on BEC (**Figure 2C**). Among participants without clinical allergy, asthma, or COPD *(n=*710), the upper BEC limit increased with age from 300 cells/μL for those aged <40 years to 400 cells/μL for subjects aged >40 years (**Table 2**). Further, excluding atopic subjects, there was a minimal change in the 95^th^ percentile compared to those without clinical allergy (**Figure 2D**). Detailed reference values of those subjects (*n*=628) are presented in **Table S2**.

**Figure 2:**
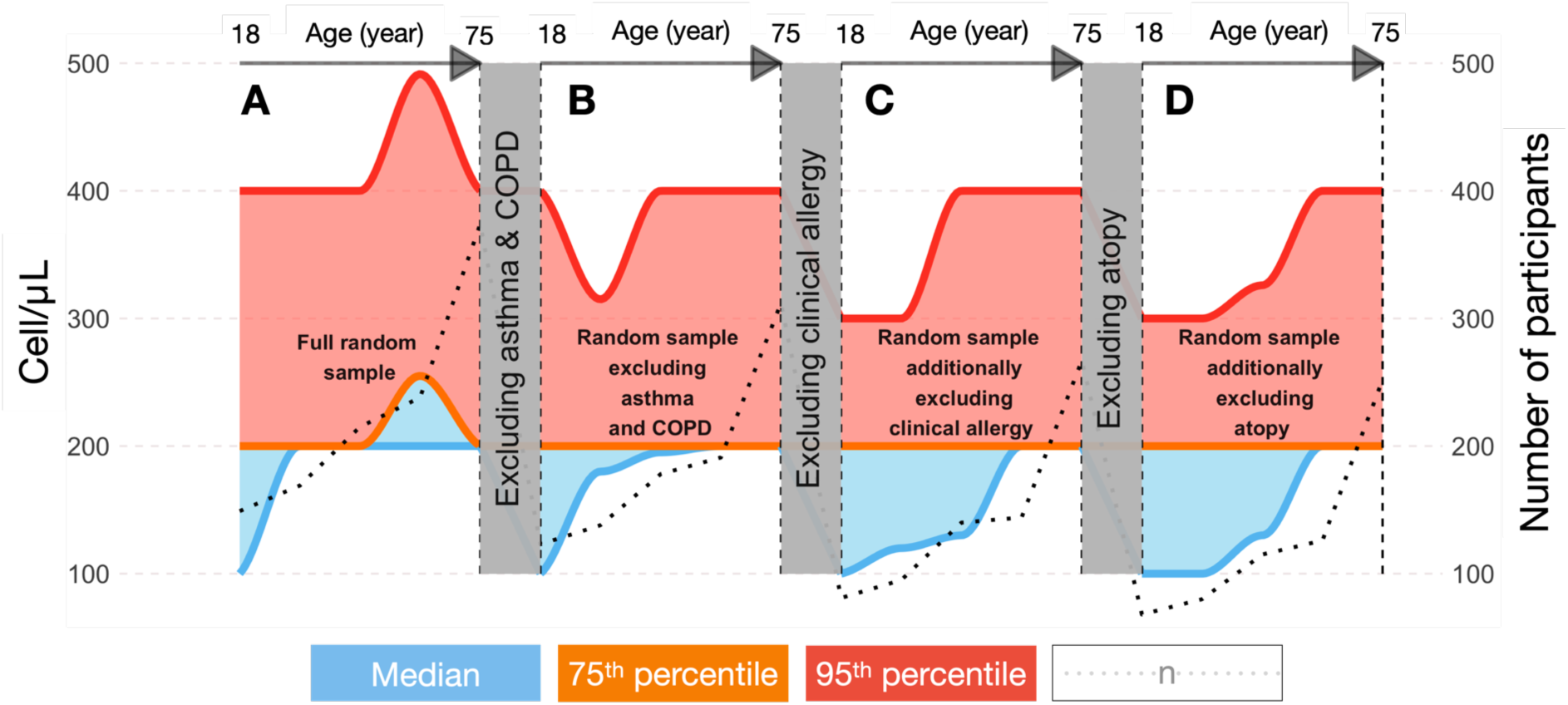
The characteristics of blood eosinophil count across different age groups, delineated for (A) the entire population-representative random sample comprising of 1,145 individuals, (B) random sample participants excluding asthma and COPD patients (*n* = 948), (C) individuals additionally devoid of clinical allergy (*n* = 710), and (D) individuals additionally devoid of atopy (*n* = 628). The depicted blue region demarcates the space between the median and 75^th^ percentile curves, while the red region signifies the interval between the 75^th^ percentile and 95^th^ percentile curves.

**Table 2.**
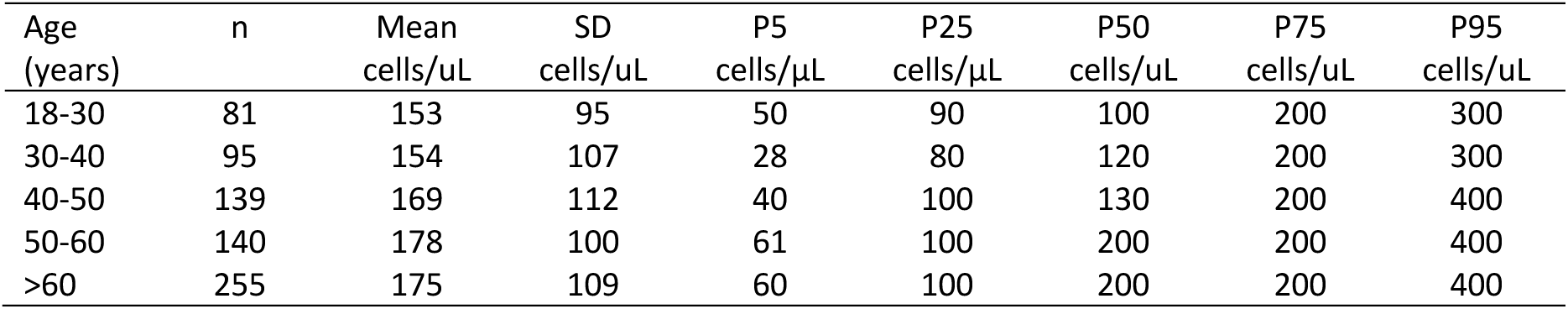
The descriptive values of BEC in non-allergic, non-asthmatic and COPD-free randomly selected participants (n = 710)

| Age (years) | n | Mean cells/uL | SD cells/uL | P5 cells/μL | P25 cells/μL | P50 cells/uL | P75 cells/uL | P95 cells/uL |
| --- | --- | --- | --- | --- | --- | --- | --- | --- |
| 18-30 | 81 | 153 | 95 | 50 | 90 | 100 | 200 | 300 |
| 30-40 | 95 | 154 | 107 | 28 | 80 | 120 | 200 | 300 |
| 40-50 | 139 | 169 | 112 | 40 | 100 | 130 | 200 | 400 |
| 50-60 | 140 | 178 | 100 | 61 | 100 | 200 | 200 | 400 |
| >60 | 255 | 175 | 109 | 60 | 100 | 200 | 200 | 400 |

The 75^th^ percentile of BEC was similar to the 95^th^ percentile in the entire random sample (**Figure 2A**). However, when excluding those with asthma, COPD, clinical allergy, or atopy, the 75^th^ percentile remained constant at 200 cells/μL across all age groups (**Figure 2B-2D**). Meanwhile, the median BEC in the full random sample increased gradually with age, from 100 cells/μL to a peak of 200 cells/μL for those aged >30 years, with similar trends in the other groups (**Figure 2**).

### The impact of asthma, COPD, and clinical allergy on BEC

To understand the rise in the upper limit of normal and the 75^th^ percentile of BEC in people aged 50-60 years in the random sample, we focused on the 95^th^ and 75^th^ percentiles of BEC in subjects with asthma, COPD, or clinical allergy. Detailed analyses of eosinophil and BEC values for the subgroups with asthma and clinical allergy in the random sample are illustrated in **Figures S1A-C**.

### Determinants of BEC

A comprehensive understanding of the determinants affecting the upper normal limit of BEC within the general population necessitates a comparative investigation of BEC upper 95^th^ percentile values across diverse demographic and clinical parameters. However, the 95^th^ percentile is a collective outcome and not individually available for each patient. Hence, to comprehend the determinants influencing BEC in the general population, we evaluated the effect of various factors, previously reported to affect BEC, on mean BEC levels in our full random sample (**Table 3**). BEC levels were significantly higher in men compared to women and in ever-smokers compared to never-smokers. BEC was significantly higher in those with than in those without clinical allergy, in those with than in those without atopy, and in those with than those without asthma. Moreover, high BEC was significantly linked to the presence of COPD, obesity, hypertension, and a minimum of one metabolic disease (**Table 3**).

**Table 3.**
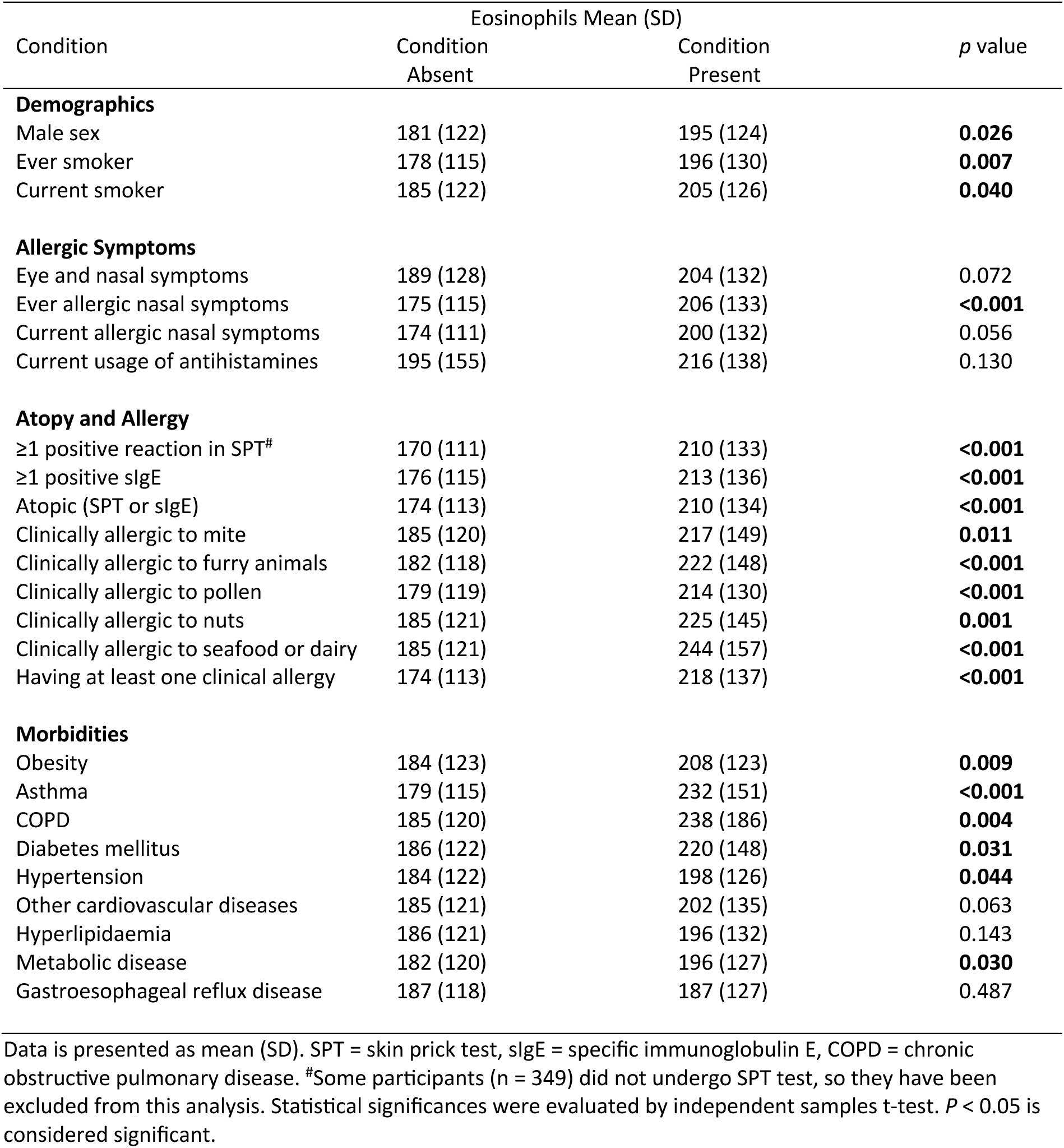
The blood eosinophil counts in the full random sample according to the demographic features and absence or presence of different conditions (N=1,145).

| Condition | Eosinophils Mean (SD) |  | p value |
| --- | --- | --- | --- |
|  | Condition Absent | Condition Present |  |
| **Demographics** |  |  |  |
| Male sex | 181 (122) | 195 (124) | **0.026** |
| Ever smoker | 178 (115) | 196 (130) | **0.007** |
| Current smoker | 185 (122) | 205 (126) | **0.040** |
| **Allergic Symptoms** |  |  |  |
| Eye and nasal symptoms | 189 (128) | 204 (132) | 0.072 |
| Ever allergic nasal symptoms | 175 (115) | 206 (133) | **<0.001** |
| Current allergic nasal symptoms | 174 (111) | 200 (132) | 0.056 |
| Current usage of antihistamines | 195 (155) | 216 (138) | 0.130 |
| **Atopy and Allergy** |  |  |  |
| ≥1 positive reaction in SPT<sup>#</sup> | 170 (111) | 210 (133) | **<0.001** |
| ≥1 positive sIgE | 176 (115) | 213 (136) | **<0.001** |
| Atopic (SPT or sIgE) | 174 (113) | 210 (134) | **<0.001** |
| Clinically allergic to mite | 185 (120) | 217 (149) | **0.011** |
| Clinically allergic to furry animals | 182 (118) | 222 (148) | **<0.001** |
| Clinically allergic to pollen | 179 (119) | 214 (130) | **<0.001** |
| Clinically allergic to nuts | 185 (121) | 225 (145) | **0.001** |
| Clinically allergic to seafood or dairy | 185 (121) | 244 (157) | **<0.001** |
| Having at least one clinical allergy | 174 (113) | 218 (137) | **<0.001** |
| **Morbidities** |  |  |  |
| Obesity | 184 (123) | 208 (123) | **0.009** |
| Asthma | 179 (115) | 232 (151) | **<0.001** |
| COPD | 185 (120) | 238 (186) | **0.004** |
| Diabetes mellitus | 186 (122) | 220 (148) | **0.031** |
| Hypertension | 184 (122) | 198 (126) | **0.044** |
| Other cardiovascular diseases | 185 (121) | 202 (135) | 0.063 |
| Hyperlipidaemia | 186 (121) | 196 (132) | 0.143 |
| Metabolic disease | 182 (120) | 196 (127) | **0.030** |
| Gastroesophageal reflux disease | 187 (118) | 187 (127) | 0.487 |

Data is presented as mean (SD). SPT = skin prick test, sIgE = specific immunoglobulin E, COPD = chronic obstructive pulmonary disease. <sup>#</sup>Some participants (n = 349) did not undergo SPT test, so they have been excluded from this analysis. Statistical significances were evaluated by independent samples t-test. *P* < 0.05 is considered significant.

### Multiple linear regression model for factors associated with higher BEC values

Multiple linear regression analyses were conducted to discern factors associated with higher BEC values considering potential confounders. To achieve this objective, two distinct analyses were performed: one including the entire random sample and another focusing solely on participants lacking any predisposing factors to eosinophilic inflammation, such as allergies, atopy, asthma, or COPD. The initial analysis revealed that only asthma and clinical allergy – not atopy– were significant determinants of BEC in the whole sample (**Table 4A**). Subsequently, to discern factors linked to higher BEC in the absence of established morbidities known to increase BEC, such as atopy, clinical allergy, asthma, and COPD, we conducted a separate analysis within this subset, in which diabetes and obesity were identified as significant predictors of BEC (**Table 4B**).

**Table 4:** Determinants of blood eosinophil count values in adjusted multiple linear regression analysis of population-representative full random sample *(N=*1,145) (A) and in participants free from asthma, COPD, clinical allergy, and atopy (*n*=628) (B).

|  | Coefficient B | Std. Error | Lower 95% CI | Upper 95% CI | P value |
| --- | --- | --- | --- | --- | --- |
| **A Full random sample** |  |  |  |  |  |
| **Demographics** |  |  |  |  |  |
| Age (years) |  |  |  |  |  |
| > 60 | 4.81 | 8.87 | -12.59 | 22.21 | 0.588 |
| < 30 | -15.97 | 11.42 | -38.37 | 6.44 | 0.162 |
| Male sex | 8.18 | 7.45 | -6.43 | 22.79 | 0.272 |
| BMI (kg/m<sup>2</sup>) |  |  |  |  |  |
| Underweight (BMI < 18.5) | -58.29 | 38.62 | -134.07 | 17.48 | 0.131 |
| Overweight (25 ≤ BMI < 30) | 4.504 | 8.30 | -11.78 | 20.79 | 0.588 |
| Obese (BMI ≥ 30) | 22.230 | 13.85 | -4.95 | 49.41 | 0.109 |
| Ever smoking | 12.308 | 8.05 | -3.49 | 28.11 | 0.127 |
| Currently smoking | 13.988 | 12.39 | -10.31 | 38.29 | 0.259 |
| **Morbidities** |  |  |  |  |  |
| Asthma | 34.09 | 10.55 | 13.39 | 54.79 | **0.001** |
| COPD | 24.60 | 21.17 | -16.95 | 66.14 | 0.246 |
| Hypertension | 12.93 | 12.62 | -11.83 | 37.69 | 0.306 |
| Diabetes | 23.14 | 19.12 | -14.38 | 60.66 | 0.226 |
| Metabolic disease (≥1) | -11.77 | 13.13 | -37.53 | 14.00 | 0.370 |
| Atopy | 7.60 | 13.31 | -18.51 | 33.71 | 0.568 |
| Clinical allergy<sup>#</sup> | 29.98 | 14.24 | 2.04 | 57.93 | **0.035** |
| **B Participants free from asthma, COPD, and clinical allergy** |  |  |  |  |  |
| **Demographics** |  |  |  |  |  |
| Age (years) |  |  |  |  |  |
| > 60 | 2.93 | 9.60 | -15.92 | 21.78 | 0.760 |
| < 30 | -0.60 | 14.24 | -28.57 | 27.37 | 0.966 |
| Male sex | 3.32 | 8.56 | -13.49 | 20.14 | 0.698 |
| BMI (kg/m<sup>2</sup>) |  |  |  |  |  |
| Underweight (BMI < 18.5) | -48.95 | 37.15 | -121.91 | 24.01 | 0.188 |
| Overweight (25 ≤ BMI < 30) | 9.120 | 9.71 | -9.95 | 28.19 | 0.348 |
| Obese (BMI ≥ 30) | 39.59 | 15.77 | 8.62 | 70.55 | **0.012** |
| Ever smoking | 12.46 | 9.03 | -5.28 | 30.21 | 0.168 |
| Currently smoking | 8.82 | 14.46 | -19.58 | 37.22 | 0.542 |
| **Morbidities** |  |  |  |  |  |
| Hypertension | 20.12 | 14.25 | -7.85 | 48.10 | 0.158 |
| Diabetes mellitus | 60.05 | 24.09 | 12.74 | 107.37 | **0.013** |
| Metabolic disease (≥1) | -22.22 | 15.02 | -51.71 | 7.27 | 0.139 |

Variables linked with the elevation of Blood Eosinophil Count (BEC) were examined through multiple linear regression analyses including all the variables presented in the table. Data are presented as the unstandardized b coefficient, standard error of b, and 95% CI. BMI= body mass index, COPD = chronic obstructive pulmonary disease.

### Binary logistic regression model for factors associated with elevated BEC values

To evaluate factors associated with BEC above the upper limit of normal (>400 cells/µl), we conducted adjusted binary logistic regression analyses similar to the above linear regressions. In the whole sample, asthma and ever smoking were significantly associated with high BEC (**Figure 3**). Diabetes, hypertension and ever smoking were significantly associated with high BEC in participants without atopy, asthma or COPD (**Figure 3**).

**Figure 3.**
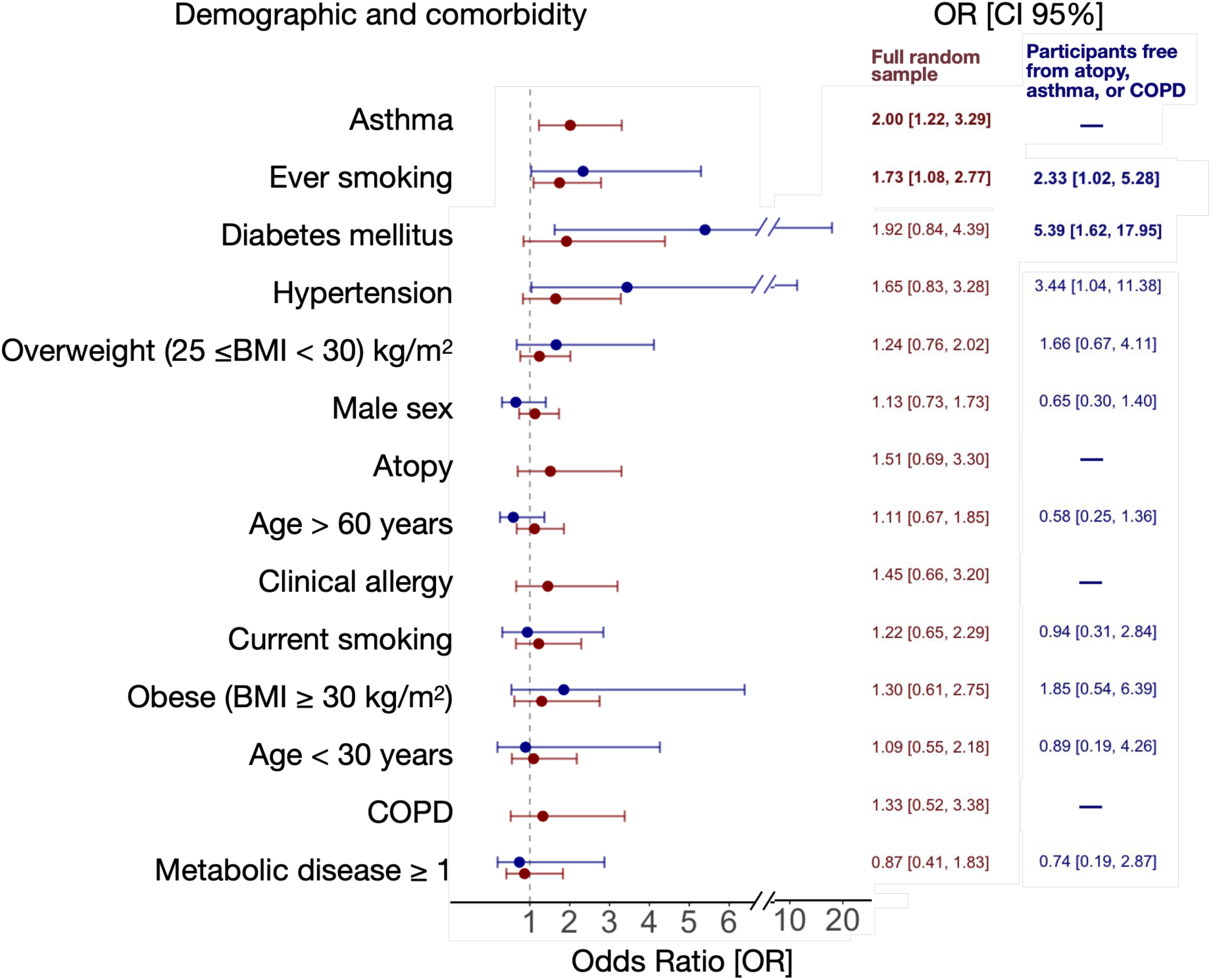
Adjusted binary logistic regression analyses (OR and 95% CI) for BEC > 400 cells/uL in the full population-representative random sample (*n* = 1145; red), and in subjects without clinical allergy, atopy, asthma, or COPD (*n* = 628; blue). Bold font indicates significant OR (*p*<0.05) associations. COPD: chronic obstructive lung disease.

## Discussion

In our population-representative random sample, the upper limit of BEC ranged from 400 to 500 cells/μL, depending on age. In individuals without asthma, COPD, or clinical allergy, the upper limit of BEC increased from 300 to 400 cells/μL after the age of 40. Sex, smoking, atopy, clinical allergy, obesity, asthma, COPD, diabetes, and hypertension were significantly associated with higher BEC levels. However, only asthma and clinical allergy in the full sample, and obesity and diabetes in healthy individuals, remained significant determinants of BEC. Asthma and ever smoking were associated with high BEC in all subjects, while diabetes, hypertension, and ever smoking were associated with high BEC in healthy participants. These findings clarify the highly needed typical BEC range in the general population, a crucial biomarker for diagnosing, treating, and managing asthma and COPD [1,2].

Several studies have attempted to establish reference values for BEC in their populations [20, 28–32]. The upper limit of normal BEC ranged from 550 to 630 cells/µL in a French population [29], from 500 to 600 cells/µL in a Moroccan population [31], and from 450 to 590 cells/µL in a Turkish population [28], depending on sex and age. In healthy individuals, the upper limit ranged from 500 to 800 cells/µL in a Thai population [30], and from 490 to 640 cells/µL in a Kenyan population [32], depending on sex. Even though the upper limit of normal BEC in our study similarly varied by age with relatively lower range, it is important to note that most of these studies do not represent the general population accurately, and their outcomes could be affected. They either analysed data from selected registries or small sample sizes instead of the large randomly selected sample from the general population. In addition, the studies using healthy populations have employed impractical strict criteria in defining their healthy population such as excluding obese, smokers, alcohol consumers, or participants with any comorbidities. In particular, our findings align more closely with those of Hartl et al. [20], who reported a 95^th^ percentile BEC range of 360 to 450 cells/µL in the general population and 290 to 350 cells/µL in healthy individuals, using a large sample size.

The reference intervals for BEC utilized in diverse contexts exhibit variability, encompassing upper limits ranging from the 75^th^ to the 90^th^, the 95^th^, and as high as the 97.5^th^ percentile [20,28–34]. Hartl et al. [20] designated the 75^th^ percentile as a threshold denoting "high blood eosinophils", employing it in their forced-entry binary multivariable logistic regression analysis. However, our analysis reveals that the sensitivity of the 75^th^ percentile to alterations is somewhat diminished, contingent upon factors such as age, atopy, clinical allergy, and smoking. Conversely, the 95^th^ percentile exhibited greater sensitivity to changes in these factors. In addition, using the 75^th^ percentile to label something as "high" implies that 25% of healthy adults are abnormal, which is not reasonable. Even those above the 95^th^ percentile do not necessarily have a disease. However, their significant deviation from the norm suggests a higher likelihood of clinically relevant abnormality. Consequently, we employed the 95^th^ percentile as the basis for our adjusted binary logistic regression analysis.

The median of BEC as reported in both healthy and general population studies exhibits a range spanning (100-200) cells/μL [8,13,14,23,35], with a proclivity to approximate 200 cells/μL in investigations with large sample sizes [8,21, 23]. Consistent with existing literature, our study observed a distribution of random sample BEC medians ranging from 100 cells/μL in the age group under 30 years to 200 cells/μL in the older age strata. Moreover, in concordance with asthma population studies, the median BEC has been reported to fluctuate between 157 and 298 cells/μL [6,7,15,16,21,36,37], demonstrating a tendency to approximate 200 cells/μL in studies involving a sizable participant pool [6,7,15,21]. In our study, the median BEC varied from 180 cells/μL among asthmatic participants under 30 years of age to 200 cells/μL in the remaining age strata. Although the absolute and average BEC values are parallel to those in other cohorts, our study uniquely helps determine the abnormality of a person’s measured value by providing reference values from a representative sample of the general population.

In our study, when comparing BEC levels between groups, we found that male sex, smoking (both current and past), atopy, clinical allergy, obesity, asthma, COPD, diabetes, and hypertension were significantly associated with higher BEC levels. Previous studies have reported similar findings regarding male sex [20,21,23,28,34], current smoking [20,34], ever smoking [34], metabolic disorders [20,23,28], and allergy parameters [20,23,34]. However, our study is the first to characterize the determinants of BEC levels using multivariate linear regression, showing that asthma and clinical allergy are significant predictors in the full random sample, while obesity and diabetes are significantly associated in healthy individuals. Giovannelli et al. [23] also identified allergic asthma, atopy, and BMI as predictors of BEC in their cohort, although BMI was only significant among non-smokers. However, there was no analysis for predictors in healthy subgroup in their study. Additionally, no prior study has explored the impact of clinical allergy on BEC within the general population; most studies have evaluated the influence of atopy. This is of particular significance given that clinical allergy retains its status as a substantive predictor of BEC even after adjustments for several confounders including atopy in our multiple linear regression analysis. Moreover, clinical allergy, rather than atopy alone, is used to define T2 inflammation in asthma guidelines [2]. The WSAS cohort presents several notable strengths. Firstly, the study encompasses a sizable sample, systematically selected without any exclusion criteria, rendering it representative of the adult population in Western Sweden. Secondly, our investigation marks the first exploration of the impact of clinical allergy, in conjunction with atopy, among the participants. Challenges often arise in diagnosing factors like clinical allergy and asthma, with many studies relying on self-reported disease. In contrast, our study employed a meticulous classification process, involving thorough clinical examinations overseen by pulmonary and allergy specialists. This approach enhances the reliability of our findings, which reveal that while asthma and clinical allergy are predictors of BEC in the general population, only asthma and a history of smoking are linked to BEC levels above the upper normal limit. The fact that allergic parameters do not pose a risk for abnormal BEC values underscores the potential utility of our established upper limit of normal for interpreting BEC values in asthma clinics. Nevertheless, it is crucial to note the limitation of the present study to adults, acknowledging that the predicted BEC values might be substantially different among children and adolescents. Additionally, our study does not account for less common infectious or chronic diseases that could potentially be linked to elevated BEC.

The upper limit of normal BEC in participants without asthma, COPD, and clinical allergy ranged between (300-400) cells/μL depending on age. GINA recommends the use of BEC to identify asthma patients with T2 inflammation using cutoff points such as 150 and 300 cells/μL [2]. Similarly, GOLD recommends the use of 300 cells/μL as a threshold of BEC to guide therapy with ICS in COPD patients [1]. It is important to note that these cutoff points fall within the normal BEC range for healthy adults in our cohort, indicating that while they are useful for predicting treatment response, they are not necessarily considered "high". However, the observation that only asthma and smoking, but not clinical allergy, are associated with abnormal BEC values highlights the potential for establishing treatment response indicators for asthma and COPD that align with reference values for healthy individuals.

In conclusion, the 95th percentile in BEC ranged depending on age between (400-500) cells/μL in a random sample of the general adult population, and between (300-400) cells/μL in individuals free from asthma, COPD, and clinical allergy. Additionally, asthma and smoking, though not allergic parameters, were identified as risk factors for having BEC above the upper limit of normal in the full sample. In subjects without asthma, clinical allergy, or COPD, smoking, diabetes, and hypertension were recognized as risk factors for elevated BEC. Consequently, these factors merit careful consideration during the clinical interpretation of BEC.

## Data Availability

All data produced in the present study are available upon reasonable request to the authors

## Abbreviations

BEC: blood eosinophil count
BMI: body mass index
COPD: chronic obstructive pulmonary disease
FEV_1_: forced expiratory volume in one second
FVC: forced vital capacity
GINA: Global Initiative for Asthma
GOLD: Global Strategy for the Diagnosis, Management and Prevention of Chronic Obstructive Lung Disease
ICS: inhaled corticosteroids
sIgE: specific immunoglobulin E
WSAS: West Sweden Asthma Study

## Authors’ contributions

Conception and design RA, SE, LL, BIN, HK,

Data analysis RA, SE, HB, BIN, HK,

Data collection RA, SE, RJ, DL, SÖ, MB, LE, MR, JL, GW, TP, BIN, HK,

Manuscript writing RA, BIN, HK

Manuscript review and editing all authors.

