## Supplementary file for "Blood Eosinophil Reference Values and Determinants in a Representative Adult Population"

**Supplementary data:**

**Methods**

**Study area and population**

The West Sweden Asthma Study (WSAS) has been described in detail previously [31]. Briefly, WSAS examines individuals aged 16 to 75 years at recruitment, randomly selected from the general population of western Sweden. Commencing in 2008, a total of 30,000 subjects within the specified age range were randomly selected through the Swedish Population Register and invited to take part in a postal survey. The sampling was stratified based on age and gender to ensure a representative reflection of the underlying population. Of the survey invited participants, excluding those untraceable (*n*=782), 18,087 individuals (response rate 62%;) participated in the study. Subsequently, a random subset of 2,000 individuals were invited to comprehensive clinical examinations, of which 1,145 (response rate 57%;) participated between 2009 and 2012 (**Figure 1**). All participants signed informed consent, and the study was approved by the regional ethics board in Gothenburg, Sweden.

**Assessment of sensitization and blood eosinophils**

Allergic sensitization was evaluated through the determination of specific Immunoglobulin E (sIgE) levels and skin prick tests for 11 aeroallergens. In brief, blood samples were obtained during clinical visits and stored at -80°C. Analysis of IgE levels against a composite of aeroallergens (Phadiatop™ [Phadia AB, Uppsala, Sweden]) was conducted, and individuals with titers of ≥0.35 kUA/L underwent additional measurements for sIgE antibody levels against specific allergens present in the mix; namely cat, dog, horse, house dust mite (*Dermatophagoides pteronyssinus*, *Dermatophagoides farinae*), mold (*Cladosporium herbarum*), birch, timothy grass, and mugwort (ImmunoCAP™ [Phadia AB, Uppsala, Sweden]), with values ≥0.35 kUA/L considered as positive. SPT involved a standard panel of 11 aeroallergens (ALK, Hørsholm, Denmark), administered after an antihistamine withdrawal period of ≥72 hours. A positive result was defined as a mean wheal diameter ≥3 mm after 15 min. Eosinophil counts were determined using standard procedures at Sahlgrenska University Hospital (Gothenburg, Sweden) with ADVIA® 2120i Hematology System (Siemens Healthineers, Erlangen, Germany), and were reported as the number of cells per microliter (μL).

**Definitions of diseases**

Clinical allergy was defined as the presence of allergic sensitization, indicated by either a positive SPT or elevated sIgE level to at least one allergen (atopy), coupled with self-reported allergic symptoms attributable to the same allergen. These included ocular, nasal, mouth, throat, skin, and gastrointestinal symptoms, as well as respiratory and asthma symptom exacerbations. Asthma was defined by any of the following criteria: 1) self-reported diagnosis of asthma by a healthcare professional or a self-reported history of asthma accompanied by respiratory symptoms or use of asthma medication in the past 12 months; 2) respiratory symptoms or use of asthma medication in the past 12 months, coupled with a positive reversibility test (≥12% and 200 ml increase in forced expiratory volume in 1 second [FEV_1_]); or 3) respiratory symptoms or use of asthma medication in the past 12 months, accompanied by a positive methacholine challenge. COPD was defined by a post-bronchodilator FEV_1_/forced vital capacity (FEV_1_/FVC) ratio <0.7, along with a smoking history of ≥10 pack-years [1]. The presence of metabolic disorder was confirmed by obesity (body mass index [BMI] ≥30 kg/m^2^) or any of the following self-reported conditions: hypertension, hyperlipidaemia, or diabetes [32].

**Results**

**Characteristics of the random sample:**

The random sample, which underwent comprehensive clinical investigation, comprised of 1,145 individuals, with 46.8% being males. The mean age of the total study sample was 50.4±15.4 years (mean ± standard deviation (SD)). Nearly half of the participants (50.8%, *n* = 582) reported no history of smoking, while 11.4% were current smokers. Notably, clinical allergy to pollen was the most common, followed by furry animals, nuts, mites, and seafood. The prevalence of asthma and gastroesophageal reflux disease was higher among participants with clinical allergy than those without clinical allergy (**Table S1)**.

**Table S1. Characteristics of WSAS I random sample subjects with or without asthma and/or clinical allergy (*N*=1,145).**

| Subjects | Without clinical allergy | | With clinical allergy | | |
| --- | --- | --- | --- | --- | --- |
|  | With asthma | Without asthma | | With asthma | Without asthma |
| *n* (%) | 65 (8.2) | 727 (91.8) | | 115 (34.1) | 222 (65.9) |
| **Demographics** |  |  | |  |  |
| Age (y) | 54.6 (15.2) | 51.8 (15.3) | | 45.2 (13.8) | 45.9 (15.1) |
| Male sex n (%) | 25 (38.5) | 324 (44.6) | | 55 (47.8) | 124 (55.9) |
| BMI, kg/m^2^ | 27.0 (4.4) | 26.0 (4.0) | | 26.8 (4.6) | 25.9 (4.1) |
| Education over 12 years, n (%) | 27 (41.5) | 318 (43.8) | | 62 (54.4) | 113 (51.1) |
| Never smokers | 26 (40.0) | 354 (48.7) | | 60 (52.2) | 136 (61.3) |
| Current smokers | 13 (20.0) | 86 (11.8) | | 14 (12.2) | 15 (6.8) |
| Smoking history (pack-years) mean (SD) | 19.4 (15.2) | 14.3 (12.7) | | 15.3 (15.8) | 12.0 (12.7) |
| **Allergic Symptoms** |  |  | |  |  |
| Age at initiation of symptoms | 23.6 (16.4) | 29.5 (18.3) | | 16.8 (14.4) | 20.1 (14.1) |
| Eyes and nasal symptoms | 20 (46.5) | 99 (31.2) | | 86 (78.2) | 148 (80.9) |
| Current allergic nasal symptoms | 42 (95.5) | 267 (82.9) | | 105 (95.5) | 180 (96.3) |
| Ever nasal polyps* | 7 (33.3) | - | | 20 (46.5) | - |
| Current usage of antihistamines | 11 (55.0) | 36 (42.9) | | 85 (91.4) | 103 (85.1) |
| **Atopy and Allergy** |  |  | |  |  |
| ≥1 positive reaction in SPT^#^ | 5 (12.5) | 55 (11.6) | | 97 (97.0) | 168 (92.3) |
| ≥1 positive sIgE | 4 (6.3) | 61 (8.7) | | 107 (95.5) | 190 (89.6) |
| Atopic (SPT or sIgE) | 7 (10.8) | 85 (11.7) | | 115 (100) | 222 (100) |
| Clinical allergy to mite | - | - | | 42 (36.5) | 42 (18.9) |
| Clinical allergy to furry animals | - | - | | 71 (61.7) | 74 (33.3) |
| Clinical allergy to pollen | - | - | | 97 (84.3) | 168 (75.7) |
| Clinical allergy to nuts | - | - | | 49 (43.8) | 40 (18.9) |
| Clinical allergy to seafood or dairy | - | - | | 19 (17) | 38 (17.9) |
| **Comorbidities** |  |  | |  |  |
| COPD | 9 (13.8) | 17 (2.3) | | 7 (6.1) | 3 (1.4) |
| Obesity (BMI ≥ 30 kg/m^2^) | 13 (20.0) | 100 (13.8) | | 26 (22.6) | 29 (13.1) |
| Diabetes mellitus | 3 (4.6) | 24 (3.3) | | 6 (5.2) | 12 (5.4) |
| Hypertension | 23 (35.4) | 174 (21.1) | | 24 (21.1) | 43 (19.5) |
| Other cardiovascular diseases | 13 (20.0) | 85 (11.7) | | 9 (7.8) | 23 (10.4) |
| Hyperlipidaemia | 15 (23.1) | 114 (15.7) | | 16 (13.9) | 31 (14.0) |
| Metabolic disease | 34 (52.3) | 287 (39.5) | | 48 (41.7) | 79 (35.6) |
| Gastroesophageal reflux disease | 31 (47.7) | 311 (42.9) | | 71 (61.7) | 92 (41.4) |

Data was presented as n (%) or mean (SD). BMI= body mass index, SPT = skin prick test, sIgE = specific immunoglobulin E, COPD = chronic obstructive pulmonary disease. ^#^Some participants (N = 349) did not undergo SPT test, so they have been excluded from the total percentage. *Nasal polyps are only evaluated among asthmatic participants (n=?).

**Blood-eosinophil count in the random sample**

**Table S2. The descriptive values of blood eosinophil count in non-atopic, non-asthmatic and COPD-free randomly selected participants according to age groups (*n* = 628)**

| Age (years) | n | Mean BEC (mean±SD)  cells/uL |  | P5  cells/uL | P25  cells/uL | P50  cells/uL | P75  cells/uL | P95  cells/uL |
| --- | --- | --- | --- | --- | --- | --- | --- | --- |
| 18-30 | 68 | 157±96 |  | 44 | 90 | 100 | 200 | 322 |
| 30-40 | 80 | 144±91 |  | 40 | 80 | 100 | 200 | 300 |
| 40-50 | 114 | 164±110 |  | 40 | 100 | 125 | 200 | 355 |
| 50-60 | 123 | 178±103 |  | 52 | 100 | 200 | 200 | 400 |
| >60 | 240 | 173±106 |  | 60 | 100 | 200 | 200 | 400 |

COPD: Chronic obstructive pulmonary disease, P5:5^th^ percentile, P25:25^th^ percentile, P50:50^th^ percentile, P75: 75^th^ percentile, P95: 95^th^ percentile, SD: standard deviation.

**The impact of asthma, COPD, and clinical allergy on BEC**

To understand the rise in the upper limit of normal and the 75^th^ percentile of BEC in people aged 50-60 years in the random sample, we focused on the 95^th^ and 75^th^ percentiles of BEC in subjects with asthma, COPD, or clinical allergy. Subjects with asthma showed an age-related increase in both the 95^th^ and 75^th^ percentiles of BEC (**Figure S1)**. To further investigate this rise among those with asthma, we analysed the complete dataset (i.e. the random sample plus an enriched asthma sample) [31]. With a larger sample of individuals with asthma (*n*=1,022), the 75^th^ percentile stabilized at 300 cells/μL across all age groups. However, the 95^th^ percentile increased in older participants with asthma (**Figure S2**). Detailed analyses of eosinophil and BEC values for the subgroups with asthma and clinical allergy in the random sample are illustrated in **Figures S1A- C**.

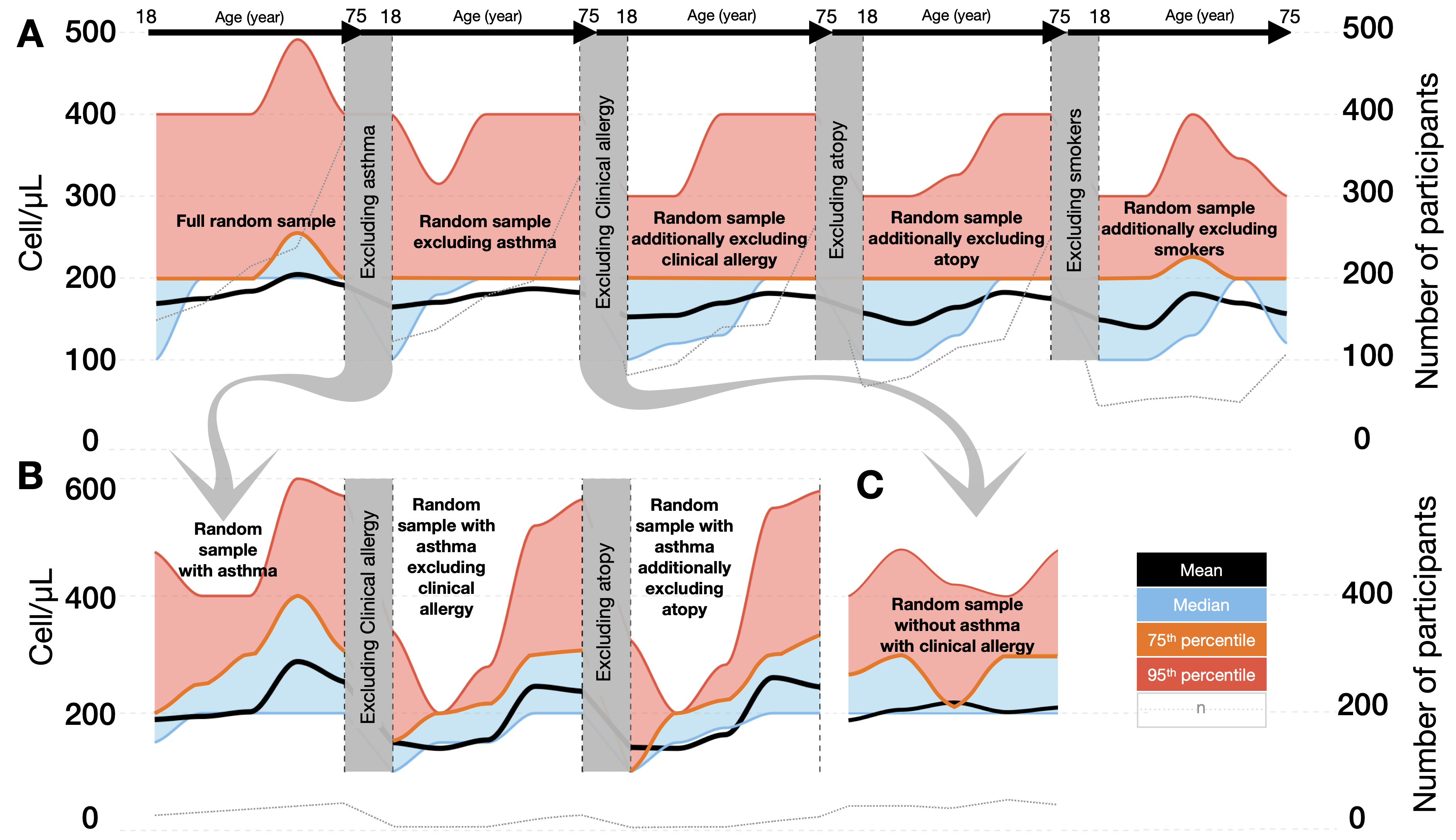

**Figure S1:** Characteristics of blood eosinophil count across different age groups, delineated for (A) the entire random sample comprising 1,145 individuals, (B) participants diagnosed with asthma (*n* = 181), and (C) individuals devoid of asthma but exhibiting clinical allergy (*n* = 222). The blue region demarcates the space between the median and 75^th^ percentile curves, while the red region signifies the interval between the 75^th^ percentile and 95^th^ percentile curves.

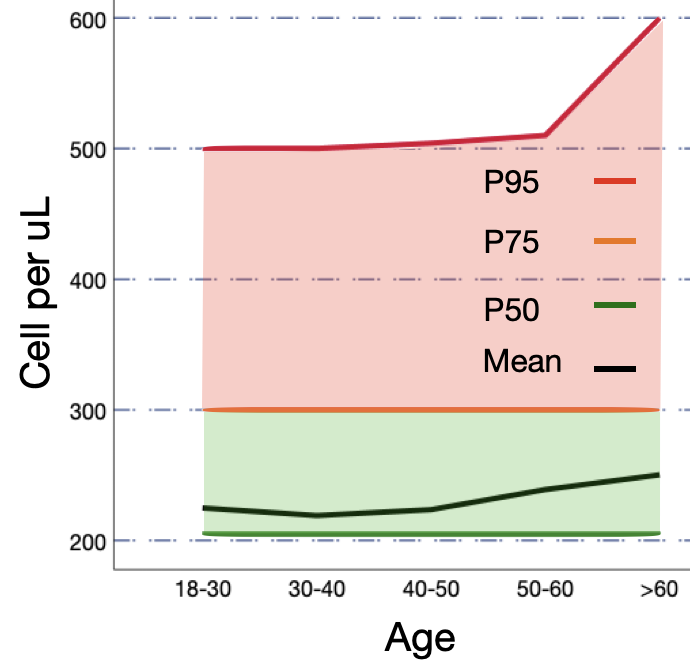

**Figure S2:** Characteristics of blood eosinophil count across different age groups, delineated for all asthma participants in WSAS, comprising 1,022 individuals. The depicted green region demarcates the space between the 50^th^ and the 75^th^ percentile curves, while the red region signifies the interval between the 75^th^ and the 95^th^ percentile curves. P50: 50^th^ percentile, P75: 75^th^ percentile, P95: 95^th^ percentile.
